# Genetic liability to psoriasis predicts severe disease outcomes

**DOI:** 10.1101/2025.03.04.25323079

**Authors:** Jake R Saklatvala, Samuel Lessard, Maris Teder-Laving, Laurent F Thomas, Ravi Ramessur, Bjørn Olav Åsvold, Anne Barton, David Baudry, John Bowes, Ben Brumpton, Vinod Chandran, Clément Chatelain, Emanuele de Rinaldis, James T Elder, David Ellinghaus, John Foerster, Andre Franke, Dafna D Gladman, Wayne Gulliver, Ulrike Hüffmeier, Laura Huilaja, Kristian Hveem, Shameer Khader, Külli Kingo, Katherine Klinger, Sulev Kõks, Wilson Liao, Rajan P Nair, Joanne Nititham, Proton Rahman, André Reis, Philip E Stuart, Kaisa Tasanen, Tanel Traks, Lam C Tsoi, Steffen Uebe, Katie Watts, BSTOP study group, Jonathan N Barker, Satveer K Mahil, Sinéad M Langan, FinnGen, Estonian Biobank research team, Sara J Brown, Mari Løset, Lavinia Paternoster, Nick Dand, Catherine H Smith, Michael A Simpson

**Author notes:** Members are listed in acknowledgements (BSTOP study group, Estonian Biobank research team). Full list of FinnGen contributors listed in Supplementary. Joint second authors (Samuel Lessard, Maris Teder-Laving, Laurent F Thomas). Joint senior authors (Nick Dand, Catherine H Smith, Michael A Simpson).

## Abstract

**Background:** Psoriasis is a common inflammatory skin disease with heterogeneous presentation. Up to 30% of individuals have severe disease with a greater surface area of skin involvement, co-morbidity burden and impact on quality of life. Prognostic biomarkers of psoriasis severity could improve allocation of clinical resources and enable earlier intervention to prevent disease progression, and a genetic biomarker would be cost-effective, stable over time, and unaffected by treatment or comorbidity.

**Methods:** Psoriasis severity was studied in four European population-based biobanks and classified based on level of clinical intervention received, with criteria for severe disease including hospitalisation due to psoriasis, use of systemic immunomodulating therapy or phototherapy. Common genetic variants, polygenic risk scores and traditional epidemiological risk factors were tested for association with severe psoriasis in each of the constituent biobanks and combined through meta-analysis. The distribution of psoriasis polygenic risk was also evaluated in a cohort of 4 151 participants in the UK-based severe psoriasis registry, BSTOP.

**Results:** In the population-based datasets, 9 738 of 44 904 individuals with psoriasis (21.7%) were classified as having severe disease. Genetic variants within the major histocompatibility complex (MHC) and the TNIP1 and IL12B psoriasis susceptibility loci were associated with severe disease at genome-wide significance (P<5.0×10^−8^). Furthermore, a strong positive correlation was observed between psoriasis susceptibility and severity effect sizes across all psoriasis susceptibility loci. An individual’s genetic liability to psoriasis as measured with a polygenic risk score (PRS) strongly associated with disease severity, with a magnitude of effect comparable to established severity risk factors such as obesity and smoking. The top 5% of psoriasis cases by genetic liability to psoriasis were 1.23-to-2.00 times as likely than the average psoriasis case to have severe disease. Psoriasis cases in the BSTOP severe disease registry were 3.10-fold enriched for a PRS that exceeded the 95th percentile established among UK Biobank psoriasis cases.

**Conclusions:** The psoriasis susceptibility PRS demonstrates utility, and may be more effective than established epidemiological factors, as a stratification tool to identify those individuals that are at greatest risk of severe disease and may benefit most from early intervention.

## Introduction

Psoriasis is a common inflammatory skin disease in which the extent of skin inflammation and impact on the affected individual can vary greatly (1). Most affected individuals have relatively mild disease, but an estimated 10-30% of individuals with psoriasis have over 10% of their body surface area affected, a commonly used benchmark for more severe skin involvement (2,3). Co-morbid burden and impact on quality of life have been shown to increase with increasing skin severity (4,5).

Many epidemiological risk factors for psoriasis have also been shown to associate with increased severity, including male sex (6), smoking (7), obesity (8), diet (9) and alcohol usage (10). Some of these risk factors are modifiable and important to proactively manage. Weight loss following lifestyle intervention, for example, has been demonstrated to reduce psoriasis severity (11). There have also been dramatic advances in therapeutics driven by better understanding of psoriasis biology, with highly targeted, though costly, systemic immunomodulators delivering clear or nearly clear skin in most individuals.

Severe psoriasis has a considerable societal impact, both directly and due to the high burden of comorbidities. The financial cost is substantial (12), including both the costs of clinical intervention and indirect costs due to lost productivity (13). Rising demand for dermatology services due to aging populations, recent targeted treatments and the increasing prevalence of common skin conditions presents a further challenge in directing care to individuals efficiently (14). The current treatment approach for psoriasis is reactive, lacking systematic approaches to stratify the patient population based on prognostic risk. Identification of reliable prognostic indicators for psoriasis would provide opportunities to ease disease burden through a stratified, proactive healthcare approach, including early onward referral from primary care settings, enhanced monitoring of highest-risk populations for skin inflammation and/or comorbidities, as well as recommendation of early lifestyle or therapeutic intervention to reduce and even prevent cumulative impact on quality of life. Psoriatic arthritis, for example, is a well-established inflammatory comorbidity of psoriasis where early identification and treatment can limit permanent functional deterioration (15). Emerging evidence in psoriasis supports the notion that early intervention may prevent inflammatory memory and increase the chance of long-term drug-free remission (16,17). For several common diseases, genetic prediction using polygenic risk scores (PRS) can identify significant proportions of individuals with increased disease risk (18), and the clinical utility of communication of genetic risk has been demonstrated in cardiovascular disease (19,20).

While psoriasis susceptibility has a strong genetic basis, with heritability of more than 60% (21), and over 100 established susceptibility loci (22) there is comparatively little evidence on the genetic involvement in disease severity. Several small studies have tested a limited number of candidate genetic variants for associations with severity (23) as assessed using objective measures of skin involvement or health care use proxies and report associations with *HLA-C*06:02* (24–27) and variation at *IL23R* (28–30), *LCE3D* (27) and *NFKBIL1* loci (29).

Here we report an analysis of 44 904 individuals with psoriasis across four large population-based datasets (UK Biobank, Trøndelag Health Study [HUNT], FinnGen and the Estonian Biobank) to systematically assess the role of genetic variation in psoriasis severity. We took advantage of wide population-scale data collection, including linked electronic health record (EHR) data, to adopt a definition of severe disease based on the level of clinical intervention received (31) and evaluated the association of genome-wide genetic variation, genetic instruments in the form of PRS and epidemiological risk factors on a dichotomised psoriasis severity phenotype.

## Methods

### Severity phenotype definition

We followed recently published guidelines (31) suggesting that a dichotomous definition of psoriasis severity could be employed in population-based datasets, using level of clinical intervention recorded as a proxy for severe disease. Individuals with psoriasis diagnoses were categorised as having severe disease if they had evidence of hospitalisation due to psoriasis or use of systemic immunomodulators (conventional systemic agents, targeted biologics, oral small molecule inhibitors) or phototherapy (narrow-band ultraviolet B radiation, psoralen with ultraviolet A radiation). Remaining individuals with psoriasis were categorised as non-severe. These criteria were applied to individuals with psoriasis in four population-based datasets – Estonian Biobank (32), FinnGen (33), HUNT (34) and UK Biobank (35), with full details of codes and data fields used in **Supplementary Data 1-6**.

### Genotyping and genetic association testing

Genome-wide association studies (GWAS) were performed separately in each cohort using logistic mixed-effect models comparing individuals with severe disease to those with non-severe disease, controlling for age, sex, genotyping batch(es), relatedness and an appropriate number of ancestry principal components (full details of genotyping, imputation, and cohort-specific models in **Supplementary Methods**).

PRS distributions (described later in **Methods**) within the UK Biobank cohort were compared to distributions of equivalent PRS in BSTOP (Biomarkers and Stratification To Optimise outcomes in Psoriasis). BSTOP is an ongoing prospective observational study of patients with a primary diagnosis of moderate‒severe plaque psoriasis across >70 UK dermatology centres, which includes biological sample collection. Full inclusion criteria have been described previously (36), including having started conventional systemic or biologic therapy within the previous 6 months. Details of genotyping are described in **Supplementary Methods**.

### QC and meta-analysis

Post-GWAS quality control and harmonisation of summary statistics were performed using GWASinspector (37), mapping to Genome Reference Consortium Human Build 37 patch release 13 reference and filtering for variants with INFO>0.7 and MAF>1%. Fixed-effects standard error-weighted meta-analysis was performed using METAL software (version release: 2020-05-05) (38).

### Genetic Correlation

Genetic correlations were calculated using LDSC (39), comparing the summary statistics of (i) the severe disease meta-analysis and (ii) a psoriasis susceptibility meta-analysis, constraining intercepts to 1. The susceptibility summary statistics were derived from a recently published meta-analysis (22), reanalysed after removing studies that overlap with the cohorts used in the present work (total 19 842 psoriasis cases and 33 108 controls, **Supplementary Data 7**).

### Effect size regression

Of the 109 loci (linkage equilibrium [LD]-partitioned genomic regions) associated with psoriasis at genome-wide significance (P<5.0×10^−8^) in the latest susceptibility meta-analysis (22), 100 retained genome-wide significant variants when considering only variants that were present in all four severe disease GWAS. For each of these 100 susceptibility loci the lead available variant was used in a regression of effect sizes between susceptibility and severe disease, using a Deming regression (constraining the intercept to zero) that accounts for measurement error in both variables. Effect size regressions were performed comparing susceptibility meta-analysis effect sizes against (a) severe disease meta-analysis effect sizes and (b) effect sizes from each severe disease GWAS individually. To check that the observed relationships were robust to the inclusion of three of our severe disease datasets (Estonian Biobank, HUNT and UK Biobank) in the susceptibility meta-analysis, the following sensitivity analyses were performed:

1. Using the same 100 lead variants but recalculating the susceptibility effect sizes in a meta-analysis that excludes our severe disease cohorts (described above and in **Supplementary Data 7**).
2. Selecting lead variants and corresponding effect sizes for 65 loci that achieve genome-wide significance in the recalculated susceptibility meta-analysis (excluding severe disease cohorts).

For each cohort, an additional follow-up sensitivity analysis was performed omitting the lead variants from the major histocompatibility complex (MHC) LD block due to its large individual effect size.

### Polygenic risk score

Variants eligible for inclusion in PRS were those available in all of (i) the latest psoriasis susceptibility meta-analysis (22) (ii) all four of the severe disease GWAS summary statistics and (iii) BSTOP imputed genotyping data. 6 461 913 variants in total were present across all datasets. Two different strategies were employed to construct psoriasis susceptibility PRS:

1. PRS_GWS_ – 65 variants: Genome-wide significant (GWS) variants in the reanalysed susceptibility meta-analysis were assigned to LD-independent blocks (40), and the lead variants from each GWS block was incorporated into the PRS and weighted according to its effect size estimate.
2. PRS_full_ – 513 461 variants: summary statistics from the reanalysed susceptibility meta-analysis (omitting Estonian Biobank, HUNT, UK Biobank and BSTOP cohorts) were analysed using the SBayesR framework to optimise weights, using the provided sparse reference LD matrix computed for 1.1 million common variants in 50 000 randomly selected, unrelated UK Biobank participants of European ancestry (41). To aid model convergence, SNPs with lower sample size (> 3 s.d.) within the susceptibility meta-analysis were omitted, as were SNPs in high LD (R^2^ > 0.9, SNP with lowest susceptibility P-value was retained). For the MHC locus (chr6, 24.0-36.3Mb), only the lead SNP (lowest P-value SNP from the LD panel: rs9380238) was included.

A sensitivity analysis was performed for both PRS_GWS_ and PRS_full_, constructed without MHC locus variants, giving a further two scores: PRS_GWS-noHLA_ and PRS_full-noHLA_.

### Epidemiological associations

Candidate epidemiological risk factors were tested for association with the severe disease phenotype in each of the four cohorts using logistic regression models. These comprised sex, age, age of psoriasis onset, smoking, alcohol intake, and various measures of adiposity (body mass index [BMI], weight, waist circumference). Epidemiological variables were defined in each cohort according to **Supplementary Data 8**. Quantitative variables were standardised to zero mean and unit variance within the psoriasis population. Effects were meta-analysed using a random-effects model (R library “metafor”).

## Results

Across four population biobanks (Estonian biobank, FinnGen, HUNT and UK Biobank), we ascertained 44 904 individuals with a diagnosis of psoriasis from EHRs and health history questionnaires (**Table 1**). This represents an average of 3.7% of participants across the four biobank studies had a linked or self-reported psoriasis diagnosis. (**Supplementary Table 1**). Within the psoriasis populations an average of 21.7% of individuals met our criteria for severe psoriatic disease (evidence of hospitalisation due to psoriasis, taking systemic immunomodulating medication or phototherapy). We noted inter-study variation in this severity proportion with FinnGen exhibiting the highest proportion of severe cases (42.2%, **Table 1**).

**Table 1:**
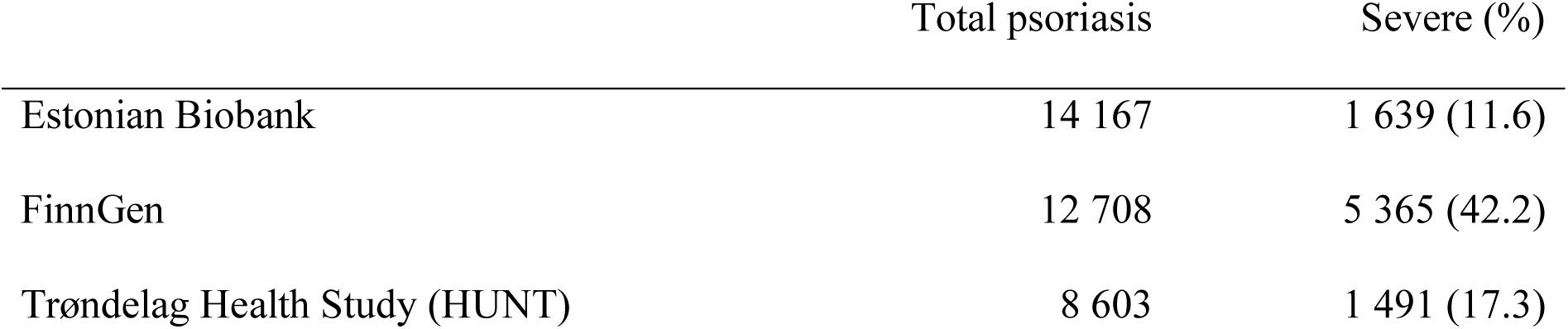
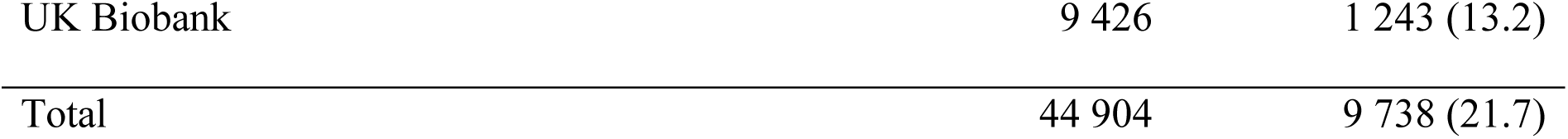
Number of individuals with severe and non-severe psoriasis within each contributing population-based cohort.

To identify genetic variation that influences an individual’s risk of developing severe psoriasis we undertook a case-control genome-wide association study in each of the four component studies. Cases were defined as individuals with a diagnosis of psoriasis and evidence of severe psoriasis (**Methods**) and controls as individuals with a diagnosis of psoriasis but with no evidence of severe disease. Association summary statistics for 6 544 261 variants tested in all four studies were combined through a fixed-effect inverse-variance weighted meta-analysis. The distribution of test statistics across genetic variants in each of the four GWAS and the resulting meta-analysis indicated that potential sources of systematic bias were adequately controlled (**Supplementary Table 1, Supplementary Figure 1-2**).

Genetic variation at three genomic loci, 6p21.33, 5q33.1 and 5q33.3, were associated with severe psoriasis with evidence of association surpassing the genome wide significance threshold (P<5.0×10^−8^, **Figure 1**). All are established psoriasis susceptibility loci and encompass the MHC region, and the candidate genes *TNIP1* and *IL12B* respectively. Among variants included in the severity meta-analysis, the lead severity-associated variant in the MHC region (rs13203895) also has the lowest susceptibility p-value (22), and the lead severity variant at 5q33.1 (rs74817271) is in near perfect LD with the lead susceptibility variant (rs8177833; R^2^ = 0.99) (42). These observations that the same genetic variation underlies susceptibility to and severity of psoriasis at both loci is consistent with the notion that there is a shared genetic component to psoriasis susceptibility and severity.

**Figure 1:**
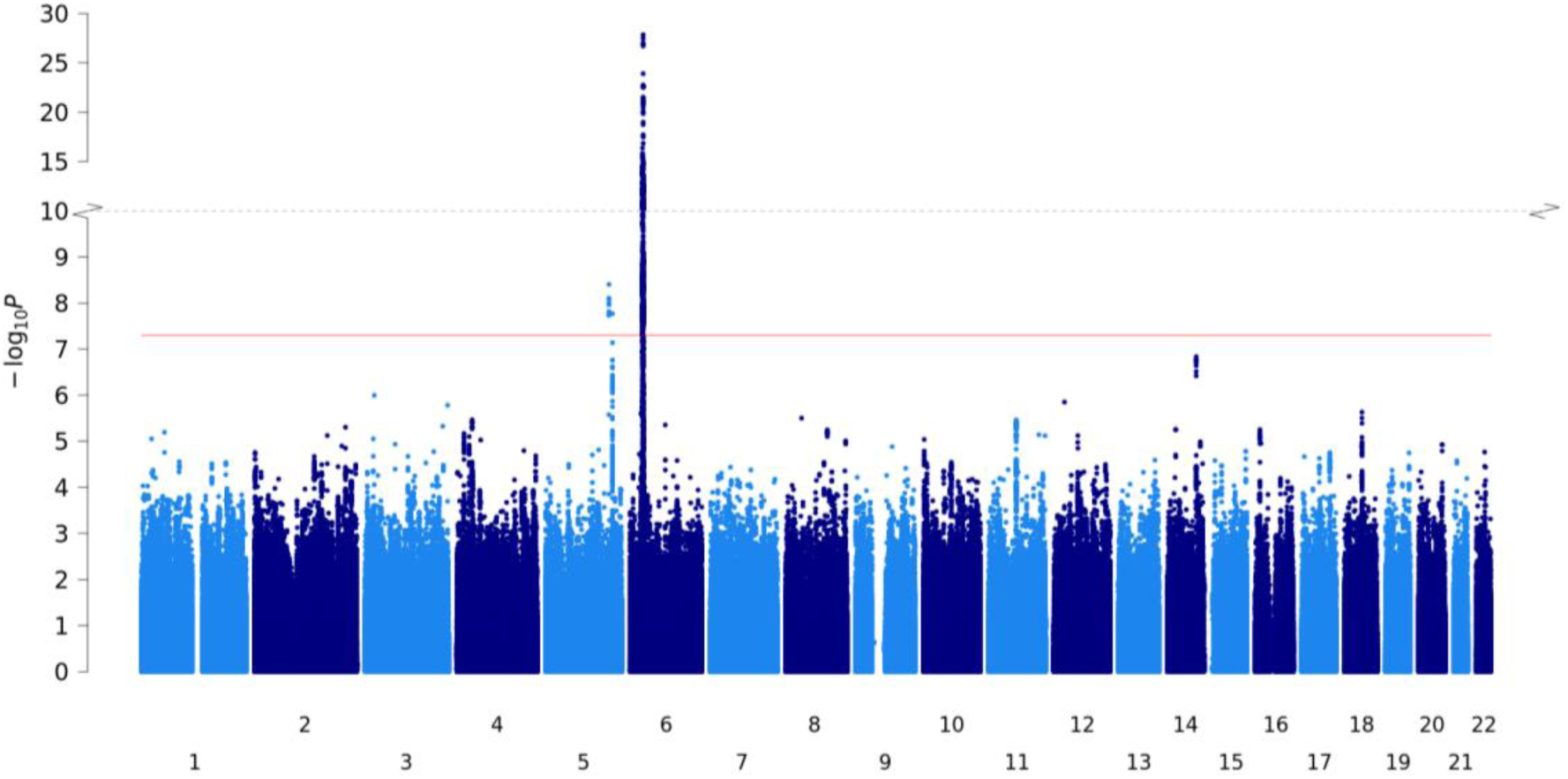
Manhattan plot for severe psoriasis GWAS meta-analysis of 4 population-based cohorts. Axes contain a point for each genetic variant (present in all datasets) ordered by chromosome and base position on the x-axis, with −log10(P-value) of association plotted on the y-axis. Red line indicates genome-wide significance threshold (P = 5 × 10^−8^).

To investigate the potential shared genetic architecture of psoriasis susceptibility and severity we evaluated the genome-wide genetic correlation between the two traits using independent datasets, revealing evidence of a substantial shared genetic component (rG = 0.697, 95% CI: 0.547-0.847). We next sought to establish whether the lead variants at 100 previously reported psoriasis risk loci are also associated with psoriasis severity. We identified 34 loci at which susceptibility alleles were nominally associated with disease severity (P<0.05) with consistent effect size direction; eight of these surpassed a Bonferroni significance threshold (P<5.0×10^−4^), including the three genome-wide significant loci. Established candidate genes at these loci include the immune-related genes *IFNLR1*, *NFKBIZ*, *IL23A*, *IL31*, *STAT2* and *NOS2* (**Supplementary Figure 3**). Across all 100 risk loci, we tested for a systematic relationship between susceptibility and severity effect sizes. This demonstrated a significant positive correlation between the reported effects on psoriasis risk and the observed effect on severe disease (β = 0.31; 95%CI: 0.23 – 0.38; **Figure 2**). This positive correlation was observed consistently across each of the four constituent studies, as well as in sensitivity analyses (**Supplementary Figure 4-5**) and is consistent with a model under which individuals with higher genetic liability to psoriasis are more likely to develop manifestations of severe disease.

**Figure 2:**
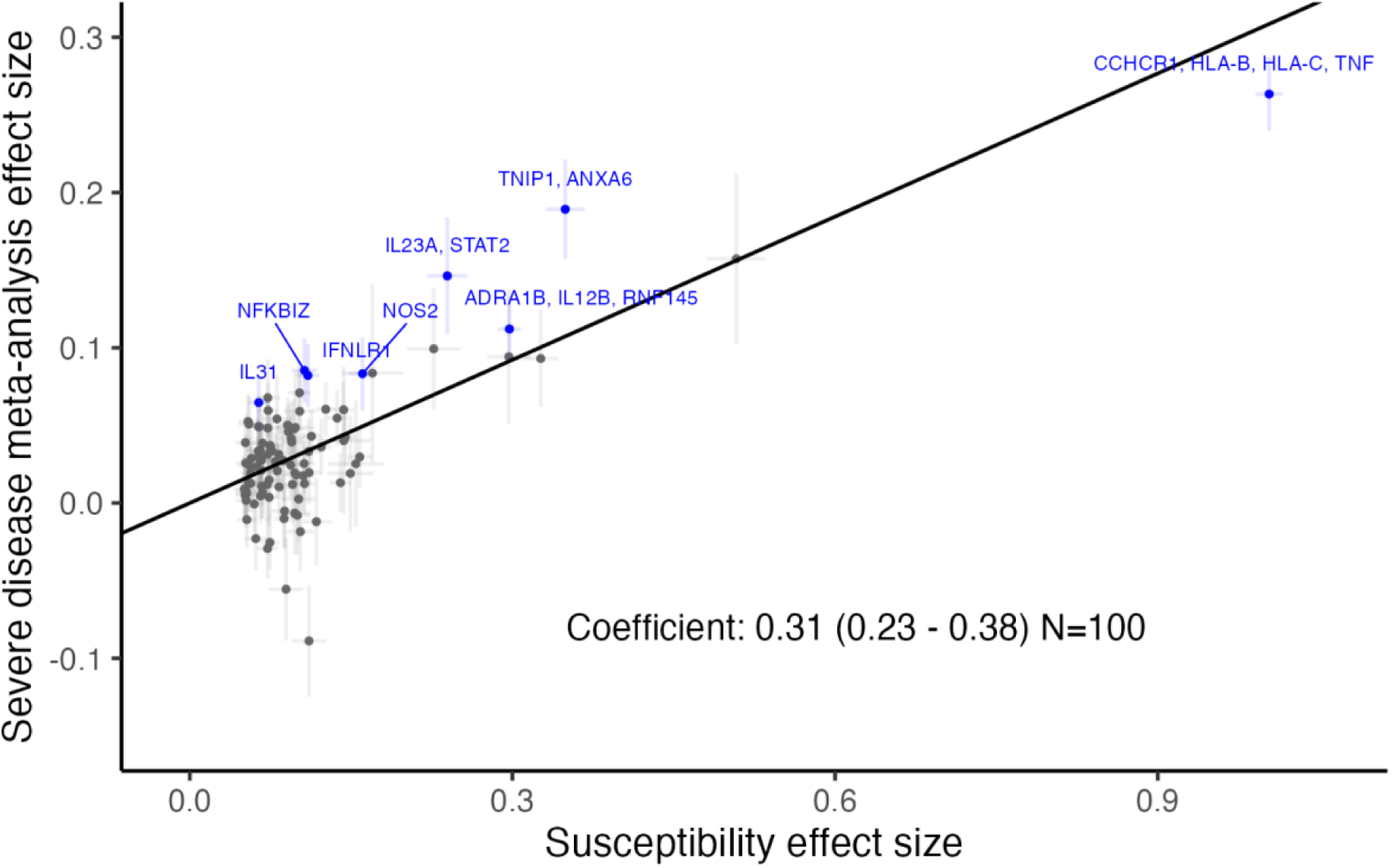
Correlation of psoriasis severity and susceptibility effects for SNPs representing 100 established susceptibility loci. x-axis: effect size (beta) for psoriasis susceptibility meta-analysis (22); y-axis: effect size (beta) estimated for severe disease in the present study. Error bars represent standard errors. Black line represents Deming regression slope fit. SNPs with Bonferroni-corrected P-values < 0.05 are highlighted blue, labelled by implicated genes (from Dand et al. 2023).

To formally investigate the hypothesis that elevated cumulative genetic liability to psoriasis predisposes individuals to develop severe disease we constructed psoriasis susceptibility PRSs and evaluated their association with psoriasis severity. PRS variant selection and weighting was undertaken using a recent large GWAS meta-analysis of psoriasis susceptibility (22), modified to exclude datasets that overlap with the current study (**Supplementary Data 7**). PRS derived from 65 genome-wide significant lead susceptibility variants (PRS_GWS_) and constructed using Bayesian multiple regression modelling (SBayesR) on genome-wide summary statistics (PRS_full_) were both significantly associated with disease severity, with effect estimates maximised using the PRS_full_ instrument (odds ratio [OR]: 1.33, 95% CI: 1.22-1.44, P = 1.01 × 10^−11^, **Figure 3**). Given the substantial effect of the MHC on psoriasis risk and the association of the same alleles with disease severity, we confirmed that both PRSs remained positively associated with disease severity after excluding genetic variation from the MHC locus (OR_full-noHLA_: 1.31, 95% CI: 1.23-1.40, P = 3.39 × 10^−16^, OR_GWS-noHLA_: 1.22, 95% CI: 1.19-1.25, P = 7.60 × 10^−55^, **Supplementary Figure 6**).

**Figure 3:**
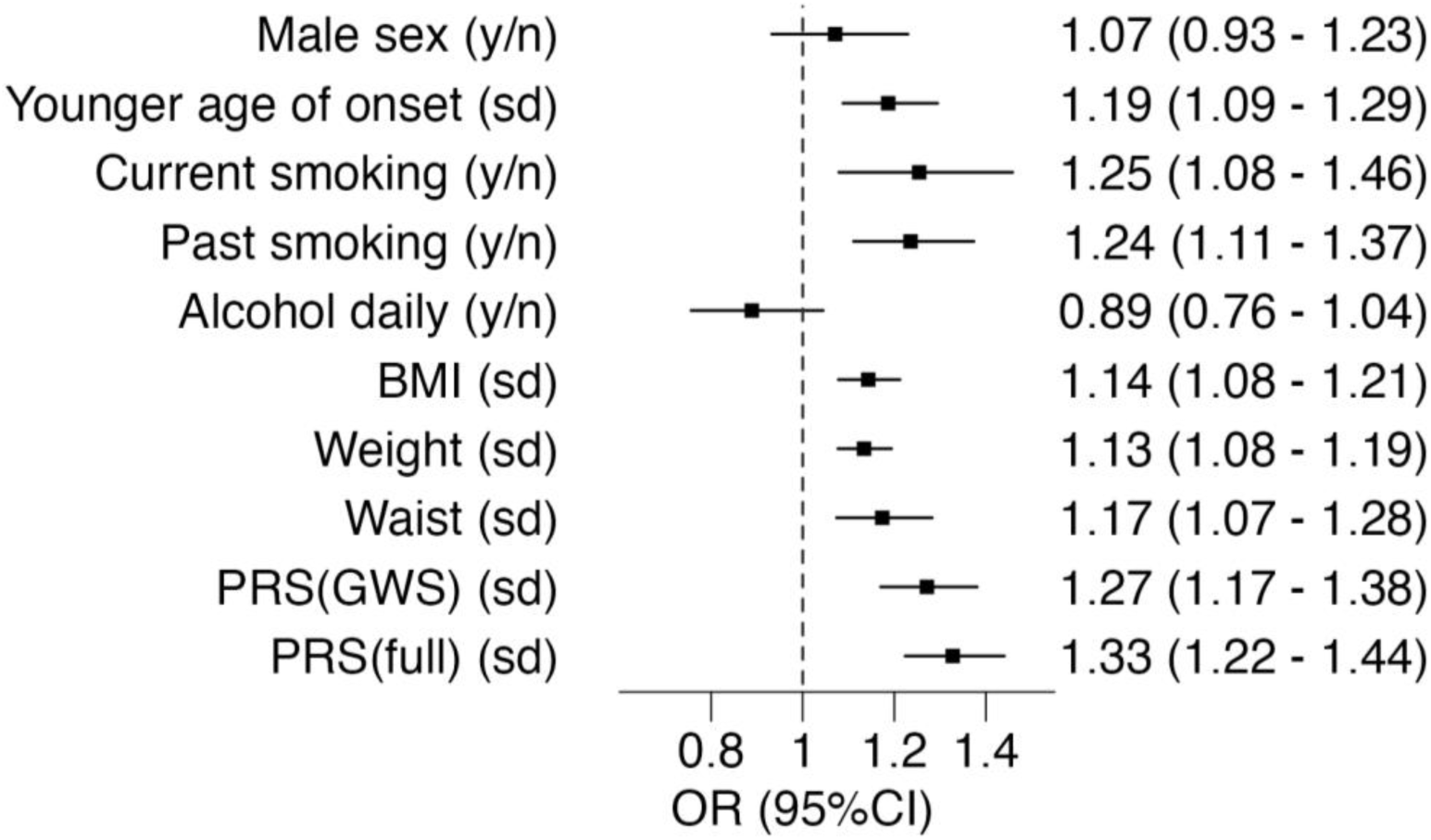
Association of epidemiological and genetic risk factors with severe psoriasis. Effect estimates are derived from a meta-analysis of unadjusted logistic regression models comparing severe to non-severe psoriasis. y/n: effect size estimated for presence of exposure (“yes”) relative to absence (”no”); sd: effect size estimated per standard deviation change in continuous exposure within the psoriasis population; OR: Odds ratio. Effect sizes presented numerically as odds ratio (with 95% confidence intervals).

To benchmark the magnitude of effect of these genetic instruments, we estimated marginal effects on severity of a series of epidemiological factors with established associations with psoriasis susceptibility and severity (**Figure 3**, **Supplementary Figure 6**). The combined sample size of 44 904 individuals represents the largest study to date of the contribution of epidemiological factors to psoriasis severity. We replicate previous reports that earlier age of onset, smoking, and higher adiposity measures (BMI, weight and waist circumference) are associated with severe disease. We saw no significant association with sex in the meta-analysis (OR: 1.07, 95% CI: 0.93-1.23, P = 0.33). Contrary to some previous reports, frequent alcohol usage was not positively associated with severe disease in any of the studies and exhibited an inverse association within UK Biobank (OR_UKBiobank_: 0.73, 95% CI: 0.62-0.85, P = 8.64 × 10^−5^).

Critically, the effect of an increase of one standard deviation of PRS_full_ on risk of severe disease is at least as strong as the effect of a binary or one standard deviation increase of any of the established epidemiological risk factors, including adiposity-related risk factors or smoking.

Given the comparative strength of association of genetic factors over established epidemiological risk factors on disease severity, we next evaluated the ability of different psoriasis susceptibility PRS thresholds to correctly classify individuals at risk of severe disease. We examined the trade-off between a PRS threshold for expedited clinical intervention and the extent to which individuals progressing to severe disease are correctly prioritised using a likelihood ratio for severe disease odds (**Supplementary Figure 7**) across the four different biobanks. As expected, there is variation between biobanks in the discriminatory ability of PRS at different thresholds. A PRS threshold at the 99^th^ percentile of polygenic risk gives a likelihood ratio of between 1.37 and 2.79 (depending on the cohort studied) but suffers from lower sensitivity compared to a more inclusive PRS threshold. A threshold at the 95^th^ percentile of genetic risk amongst the population with psoriasis offers a balance in sensitivity with an increased likelihood of developing severe disease of between 1.23 and 2.00 compared to the odds at the mean of the PRS distribution (**Figure 4, Supplementary Figure 8**).

**Figure 4:**
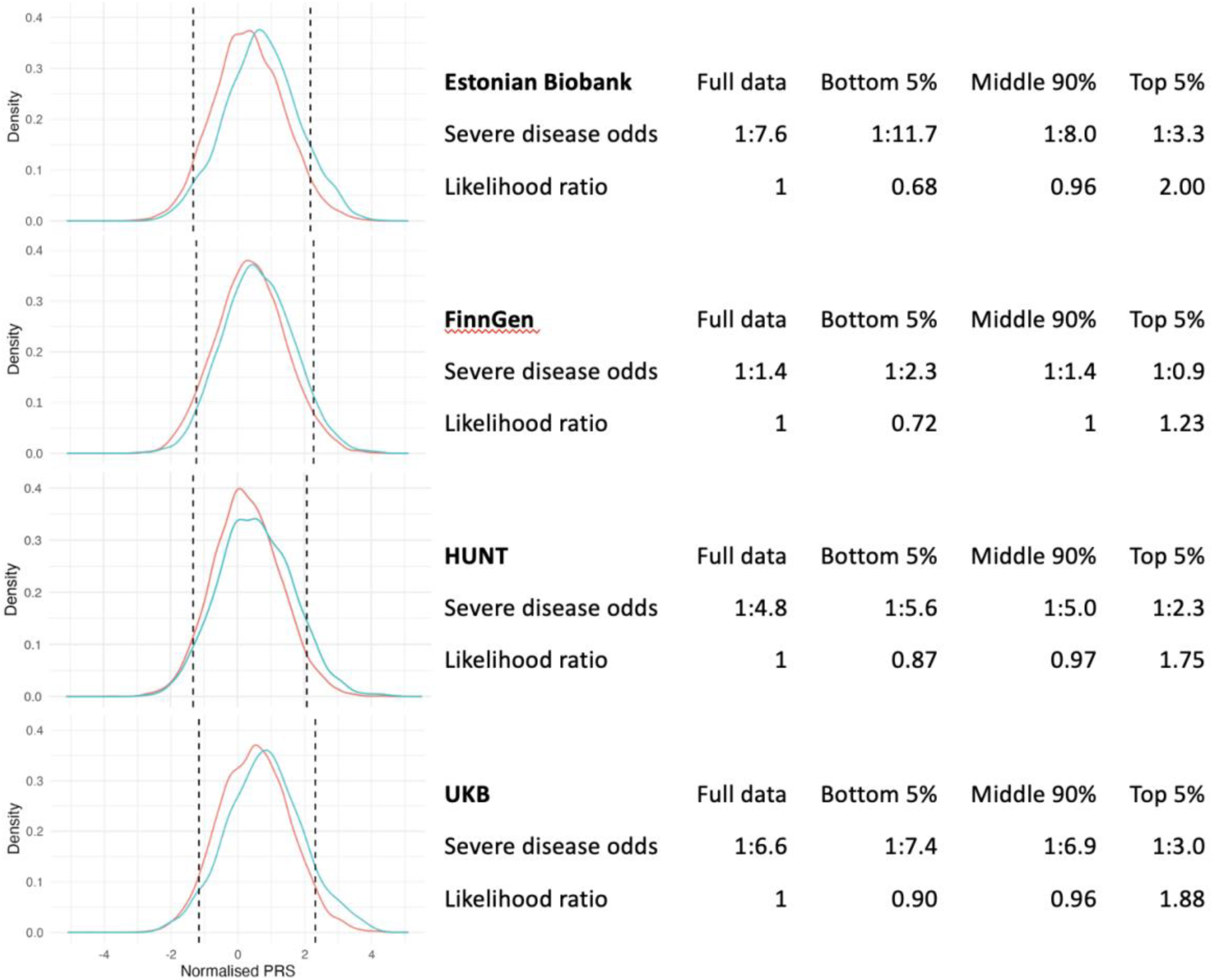
Distributions and performance of susceptibility PRS_full_ in predicting severe psoriasis cases within the cohort psoriasis population in each biobank study. Cut-offs (dashed lines) displayed for individuals within the top 5%, middle 90% and bottom 5% of the (within dataset) PRS distribution. Red line indicates PRS distribution of non-severe psoriasis population. Blue line indicates PRS distribution of severe psoriasis population. *Severe disease odds*: ratio of individuals with severe disease to individuals without severe disease. *Likelihood ratio*: ratio between the severe disease odds in each PRS group and the severe disease odds for all psoriasis cases.

To further validate the correlation of the psoriasis susceptibility PRS with disease severity, we calculated susceptibility PRS for 4 151 individuals with severe psoriasis from the BSTOP registry, a UK-based cohort of individuals ascertained from a severe psoriasis population. The mean of the distribution of the psoriasis susceptibility PRS in this group is higher than that observed among the 1 243 severe psoriasis cases in UK Biobank (P = 1.67 × 10^−37^; **Supplementary Figure 9**). Furthermore, 15.5% of the individuals in the BSTOP cohort have a PRS_full_ in excess of a threshold defined as the 95^th^ percentile of risk in the UK Biobank psoriasis population (a 3.10-fold enrichment), compared to 9.4% of severe psoriasis cases in UK Biobank (**Supplementary Table 2**).

## Discussion

The path of disease progression is a critical aspect of the psoriasis phenotype that encompasses both the severity of symptoms and the associated comorbidities, significantly affecting patients’ quality of life. Accurate prediction of who is at risk of severe disease has the potential to ensure timely and targeted interventions, to prevent long-term complications (such as psoriatic arthritis) and optimise outcomes. Our results demonstrate the potential of genetic variation in the prediction of severe psoriasis, specifically illustrating how knowledge of the genetic architecture of psoriasis susceptibility can be leveraged to predict severity.

We defined individuals with severe psoriasis based on the extent of clinical care they had received across four population biobanks. This ascertainment approach has a limitation in that differing healthcare and treatment access across different countries, as well as the extent of data capture and linkage, could introduce subtle differences in the psoriasis phenotype being represented here as severe disease. Though we observe an overall rate of severe disease that is consistent with previous studies based on body surface area (2,3), we do observe an elevated rate of severe disease based on our criteria in FinnGen, and this is likely to also be affected by the hospital-based recruitment strategy of this resource (33).

We identified three individual loci at which genetic variation was associated with psoriasis severity with genome-wide significant evidence. The association signals observed at 6p21.33, 5q33.1 and 5q33.3 are all previously established psoriasis susceptibility loci. Notably, there is also strong evidence of genetic correlation between psoriasis susceptibility and severity when considering either established psoriasis risk loci or common variation across the genome. This relationship between genetic susceptibility and severity is consistent with the liability threshold model of disease, which posits that the manifestation of a complex disease occurs when an individual’s cumulative genetic and environmental risk factors exceed a certain threshold. The results presented in this study are consistent with an extension to this model whereby, once the threshold for disease onset is surpassed, the extent of an individual’s genetic contribution to their disease liability further influences the severity of their disease. This framework enables disease severity to be conceptualised as a continuum, influenced by the extent of an individual’s genetic and environmental liability beyond the initial threshold necessary for disease manifestation.

Consistent with this model, a psoriasis susceptibility PRS is associated with disease severity. The magnitude of effect that is at least as large as established epidemiological factors that have an established association with severity (7,8). This includes BMI, weight and waist circumference and daily smoking, which are often cited as principal drivers of severe disease. We did not observe an association between daily alcohol intake and severe disease. There is conflicting evidence of the relationship between alcohol intake and severe psoriasis, with some studies showing increased intake in severe disease (43,44) and others showing decreased intake (45,46). While cross-sectional studies have limited ability to determine causal relationships, we note also that our phenotype definition may impact our ability to accurately estimate the true association: alcohol is contraindicated with use of methotrexate, one of the systemic treatments used in our severe psoriasis definition.

A clear potential use of a prognostic psoriasis severity biomarker would be the identification of individuals at initial diagnosis that are at highest risk of progressing to severe disease, and therefore most likely to benefit from early onward referral or intervention. Our analysis of different PRS thresholds highlights the challenges of balancing sensitivity and specificity. Any strata of individuals at the high end of the distribution will result in imperfect sensitivity (**Supplementary Figure 7**). The predictive performance varied across cohorts, and the discriminatory ability was most limited in FinnGen, in which a higher proportion of individuals were defined as having severe disease (**Supplementary Table 1**).

Whilst we explored the predictive ability of a genetic instrument on its own, we expect the discriminatory ability to be enhanced through the incorporation of epidemiological factors as well as typing at psoriasis presentation, which are not available in the datasets studied here. Fine-tuning the optimal proportion of high-risk individuals to monitor will require assessment of sensitivity and specificity alongside the economic impact of intervention. A further consideration will be the views of individuals living with psoriasis as to what level of risk is appropriate to trigger early interventions. Ultimately prospective studies will be required to establish the utility of severity prediction and stratification using PRS, whether this improves upon the current paradigm of treating severe disease reactively or stratifying using epidemiological factors alone.

We note the potential impact on our findings of the misclassification of unaffected individuals as psoriasis cases, which would be expected to disproportionately occur in the non-severe group (since the additional evidence that we require to define a psoriasis case as severe also corroborates the psoriasis diagnosis). However, evidence suggests that non-specialist diagnosis of psoriasis tends to be accurate, with at least 90% of psoriasis primary care diagnosis codes corroborated by GPs (47), and that self-reported psoriasis is a good proxy for physician-diagnosed disease (48).

The time taken to progress to severe disease is an important component of disease progression not assessed in the current study and critical to understand when considering future clinical utility of any prognostic strategy. Such an analysis is challenging with current data resources as severity is a composite measure spanning many different sources (across different cohorts). Therefore, defining the point in time when initial diagnosis was made and when progression to severe disease is achieved is limited by the completeness of each individual’s EHR. More complete datasets will be required to investigate the relationship between disease risk burden and the rate of progression to severe disease.

## Conclusions

In summary, our study illustrates that a psoriasis susceptibility PRS can predict disease severity, with effect sizes comparable to established epidemiological risk factors such as BMI and smoking. These insights pave the way for a more stratified and proactive healthcare approach, where early identification of high-risk individuals could lead to timely interventions, potentially mitigating the long-term impact of severe psoriasis and improving patient outcomes.

## List of abbreviations

BMI: body mass index
BSA: Body surface area
BSTOP: Biomarkers and Stratification To Optimise outcomes in Psoriasis
EHR: electronic health record
GWAS: Genome-wide association study
GWS: Genome-wide significant
HUNT: Trøndelag Health Study
LD: linkage equilibrium
OR: odds ratio
PRS: polygenic risk score

## Declarations

### Ethics approval and consent to participate

As required by the Estonian Human Genes Research Act, all participants joining the Estonian Biobank have signed an informed consent form to ensure voluntary and informed participation.

Participants in FinnGen provided informed consent for biobank research on basis of the Finnish Biobank Act.

Participation in HUNT is based on informed consent, and the study has been approved by the Norwegian Data Protection Authority and the Regional Committee for Medical and Health Research Ethics in Central Norway (REK Reference number 27420).

The UK Biobank study was approved by the National Health Service National Research Ethics Service (ref. 11/NW/0382), and all participants provided written informed consent to participate in the UK Biobank study. Information about ethics oversight in the UK Biobank can be found at https://www.ukbiobank.ac.uk/ethics/.

BSTOP is an ongoing prospective observational study of patients with moderate‒severe plaque psoriasis across >70 UK dermatology centres, which includes biological sample collection. Participants were adults (aged >16 years) at the time of recruitment; and provided written informed consent (BSTOP received approval from the London – Westminster Research Ethics Committee [previously called South East London REC 2 when originally approved in 2011], reference 11/H0802/7).

### Consent for publication

Not applicable

### Availability of data and materials

- Severe disease GWAS meta-analysis summary statistics will be deposited to GWAScatalog and will be publicly available as of the date of publication. The psoriasis polygenic risk score weights will be deposited to PGScatalog and will be publicly available as of the date of publication.
- Estonian Biobank: Estonian Biobank is open to researchers worldwide with clear standard operating procedures for data access (https://genomics.ut.ee/en/content/estonian-biobank).
- FinnGen: Based on National and European regulations (GDPR) access to individual-level sensitive health data must be approved by national authorities for specific research projects and for specifically listed and approved researchers. The health data described here was generated and provided by the National Health Register Authorities (Finnish Institute of Health and Welfare, Statistics Finland, KELA, Digital and Population Data Services Agency) and approved, either by the individual authorities or by the Finnish Data Authority, Findata, for use in the FinnGen project. Therefore, we, the authors of this paper, are not in a position to grant access to individual-level data to others. However, any researcher can apply for the health register data from the Finnish Data Authority Findata (https://findata.fi/en/permits/) and for individual-level genotype data from Finnish biobanks via the Fingenious portal (https://site.fingenious.fi/en/) hosted by the Finnish Biobank Cooperative FINBB (https://finbb.fi/en/). All Finnish biobanks can provide access for research projects within the scope regulated by the Finnish Biobank Act, which is research utilizing the biobank samples or data for the purposes of promoting health, understanding the mechanisms of disease or developing products and treatment practices used in health and medical care.
- HUNT: The HUNT data reported in this study cannot be deposited in a public repository because it is governed by Norwegian law. To request access, researchers associated with Norwegian research institutes can apply for the use of HUNT data and samples with approval by the Regional Committee for Medical and Health Research Ethics. Researchers from other countries may apply if collaborating with a Norwegian Principal Investigator. Information for data access can be found at https://www.ntnu.edu/hunt/data. The HUNT variables are available for browsing on the HUNT databank at https://hunt-db.medisin.ntnu.no/hunt-db/. Use of the full genetic dataset requires the use of an approved secure computing solution such as the HUNT Cloud (https://docs.hdc.ntnu.no). Data linkages between HUNT and health or administrative registries require that the principal investigator has obtained project-specific approval for such linkage from the Regional Committee for Medical and Health Research Ethics, Norway and each registry owner.
- UK Biobank: The UK Biobank resource is available to bona fide researchers for health-related research in the public interest (https://www.ukbiobank.ac.uk/enable-your-research).
- Biomarkers of Systemic Treatment Outcomes in Psoriasis data are available for approved research use by making an application to the BSTOP Data Access Committee (https://www.kcl.ac.uk/lsm/research/divisions/gmm/departments/dermatology/rese arch/stru/groups/bstop/documents).

### Competing interests

CHS is an investigator on EC-IMI funded consortia with multiple industry partners (see HIPPOCRATES-IMI.eu and BIOMAP-IMI.eu); departmental research funding from Sanger as part of Open Targets programme, Astrazeneca, Boehringer Ingelheim.

SJB has received research funding (but no personal financial benefits) from the Wellcome Trust (senior research fellowship ref 220875/Z/20/Z), UKRI, Medical Research Council, Rosetrees Trust, Stoneygates Trust, British Skin Foundation, Charles Wolfson Charitable Trust, anonymous donations from people with eczema, Unilever, Pfizer, Abbvie, Sosei-Heptares, Janssen, European Lead Factory (multiple industry partners) and the BIOMAP consortium (EC-IMI project ref 821511).

SL, CC, SK, EdR, KK are Sanofi employees and may hold shares and/or stock options in the company.

SKM reports departmental income from AbbVie, Almirall, Eli Lilly, Janssen, Leo, Novartis, Pfizer, Sanofi, and UCB, outside the submitted work.

DDG has received grants and/or consulting fees from Abbvie, Amgen, BMS, Eli Lilly, Janssen, Novartis, Pfizer and UCB.

No other authors declare a conflict of interest.

### Authors’ contributions (Authors with identical initials are enumerated according to position in author list)

Conceptualization: JRS, RR, JNB, SML, SJB, LP, ND, CHS, MAS

Methodology: JRS, KW, LP, ND, CHS, MAS

Validation: SL, MTL, LFT, BB, CC, EdR, LH, KH, SK1, KK1, KK2, KT, ML

Formal analysis: JRS, SL, MTL, LFT

Investigation: JRS, SL, MTL, LFT, RR, BOÅ, BB, CC, EdR, LH, KH, SK1, KK1, KK2, KT, KW, SKM, SML, SJB, ML, LP, ND, CHS, MAS

Resources: SL, MTL, LFT, BOÅ, AB, DB, JB, BB, VC, CC, EdR, JTE, DE, JF, AF, DDG, WG, UH, LH, KH, SK1, KK1, KK2, SK2, WL, RPN, JN, PR, AR, PES, KT, TT, LCT, SU, JNB, ML, LP, ND, CHS, MAS, FinnGen, BSTOP study group, Estonian Biobank research team

Data Curation: JRS, SL, MTL, LFT, RR, BOÅ, BB, CC, EdR, LH, KH, SK1, KK1, KK2, KT, KW, SKM, SML, SJB, ML, LP, ND, CHS, MAS

Writing - Original Draft: JRS, ND, CHS, MAS

Writing - Review & Editing: All authors

Visualization: JRS, SL, MTL, LFT

Supervision: ML, LP, ND, CHS, MAS

Project administration: LP, ND, CHS, MAS

Funding acquisition: JNB, ND, CHS, MAS

## Acknowledgements

This project has received funding from the Innovative Medicines Initiative 2 Joint Undertaking (JU) under grant agreement number 821511 (Biomarkers in Atopic Dermatitis and Psoriasis). The JU receives support from the European Union’s Horizon 2020 research and innovation programme and the European Federation of Pharmaceutical Industries and Associations. This publication reflects only the author’s view and the JU is not responsible for any use that may be made of the information it contains.

This research was conducted using the UK Biobank resource under application no. 15147 and 81499. Linked health data copyright © (2023), NHS England. Re-used with the permission of the NHS England and UK Biobank. All rights reserved.

Support for the study was received from the Department of Health through the National Institute for Health Research (NIHR) BioResource Clinical Research Facility and comprehensive Biomedical Research Centre awards to Guy’s and St Thomas’ National Health Service Foundation Trust in partner hip with King’s College London and King’s College Hospital National Health Service Foundation Trust (reference: BRC_1215_20006).

We would like to thank the Psoriasis Association for ongoing support and funding since the inception of Biomarkers of Systemic Treatment Outcomes in Psoriasis (reference: RG2/10: RG2/10). The authors acknowledge the invaluable support of the NIHR through the clinical research networks and its contribution in facilitating recruitment to both Biomarkers of Systemic Treatment Outcomes in Psoriasis and the British Association of Dermatologists Biologics and Immunomodulators Register.

Members of the BSTOP Study Group (excluding individually named authors of this work) are Nadia Aldoori, Mahmud Ali, Alex Anstey, Fiona Antony, Charles Archer, Suzanna August, Periasamy Balasubramaniam, Kay Baxter, Anthony Bewley, Alexandra Bonsall, Victoria Brown, Katya Burova, Aamir Butt, Mel Caswell, Sandeep Cliff, Mihaela Costache, Sharmela Darne, Emily Davies, Claudia DeGiovanni, Trupti Desai, Bernadette DeSilva, Victoria Diba, Eva Domanne, Harvey Dymond, Caoimhe Fahy, Leila Ferguson, Maria-Angeliki Gkini, Alison Godwin, Fiona Hammonds, Sarah Johnson, Teresa Joseph, Manju Kalavala, Mohsen Khorshid, Liberta Labinoti, Nicole Lawson, Alison Layton, Tara Lees, Nick Levell, Helen Lewis, Calum Lyon, Sandy McBride, Sally McCormack, Kevin McKenna, Serap Mellor, Ruth Murphy, Paul Norris, Caroline Owen, Urvi Popli, Gay Perera, Nabil Ponnambath, Helen Ramsay, Aruni Ranasinghe, Saskia Reeken, Rebecca Rose, Rada Rotarescu, Ingrid Salvary, Kathy Sands, Tapati Sinha, Simina Stefanescu, Kavitha Sundararaj, Kathy Taghipour, Michelle Taylor, Michelle Thomson, Joanne Topliffe, Roberto Verdolini, Rachel Wachsmuth, Martin Wade, Shyamal Wahie, Sarah Walsh, Shernaz Walton, Louise Wilcox, and Andrew Wright.

We want to acknowledge the participants and investigators of FinnGen study. The full list of FinnGen contributors are provided in **Supplementary Data 9**. The FinnGen project is funded by two grants from Business Finland (HUS 4685/31/2016 and UH 4386/31/2016) and the following industry partners: AbbVie Inc., AstraZeneca UK Ltd, Biogen MA Inc., Bristol Myers Squibb (and Celgene Corporation & Celgene International II Sàrl), Genentech Inc., Merck Sharp & Dohme LCC, Pfizer Inc., GlaxoSmithKline Intellectual Property Development Ltd., Sanofi US Services Inc., Maze Therapeutics Inc., Janssen Biotech Inc, Novartis Pharma AG, and Boehringer Ingelheim International GmbH. Following biobanks are acknowledged for delivering biobank samples to FinnGen: Auria Biobank (www.auria.fi/biopankki), THL Biobank (www.thl.fi/biobank), Helsinki Biobank (www.helsinginbiopankki.fi), Biobank Borealis of Northern Finland (https://www.ppshp.fi/Tutkimus-ja-opetus/Biopankki/Pages/Biobank-Borealis-briefly-in-English.aspx), Finnish Clinical Biobank Tampere (www.tays.fi/en-US/Research_and_development/Finnish_Clinical_Biobank_Tampere), Biobank of Eastern Finland (www.ita-suomenbiopankki.fi/en), Central Finland Biobank (www.ksshp.fi/fi-FI/Potilaalle/Biopankki), Finnish Red Cross Blood Service Biobank (www.veripalvelu.fi/verenluovutus/biopankkitoiminta), Terveystalo Biobank (www.terveystalo.com/fi/Yritystietoa/Terveystalo-Biopankki/Biopankki/) and Arctic Biobank (https://www.oulu.fi/en/university/faculties-and-units/faculty-medicine/northern-finland-birth-cohorts-and-arctic-biobank). All Finnish Biobanks are members of BBMRI.fi infrastructure (www.bbmri.fi). Finnish Biobank Cooperative - FINBB (https://finbb.fi/) is the coordinator of BBMRI-ERIC operations in Finland. The Finnish biobank data can be accessed through the Fingenious® services (https://site.fingenious.fi/en/) managed by FINBB.

The Trøndelag Health Study (The HUNT Study) is a collaboration between HUNT Research Centre (Faculty of Medicine and Health Sciences, NTNU, Norwegian University of Science and Technology), Trøndelag County Council, Central Norway Regional Health Authority, and the Norwegian Institute of Public Health. The genotyping in HUNT was financed by the National Institutes of Health; University of Michigan; the Research Council of Norway; the Liaison Committee for Education, Research and Innovation in Central Norway; and the Joint Research Committee between St Olavs hospital and the Faculty of Medicine and Health Sciences, NTNU. The genetic investigations of the HUNT Study are a collaboration between researchers from the HUNT Center for Molecular and Clinical Epidemiology (formerly known as the K.G. Jebsen Center for Genetic Epidemiology as of August 1[st] 2023), NTNU, and the University of Michigan Medical School and the University of Michigan School of Public Health. We thank HUNT participants for donating their time, samples, and information to help others; clinicians and other employees at Nord-Trøndelag Hospital Trust for their support and for contributing to data collection.

