## Supplementary Information for "Genetic liability to psoriasis predicts severe disease outcomes"

### Supplementary material
### for

### Genetic liability to psoriasis predicts severe disease outcomes

**Supplementary Table 1**: Number of individuals with severe and any psoriasis within each contributing GWAS cohort and the meta-analysis, along with summary data on number of high-quality (HQ) variants included (autosomal, INFO > 0.7 and MAF > 1% for contributing cohorts, HQ variants present in all four cohorts for the meta-analysis) and genomic inflation statistic ($\lambda$) for the GWAS.

| Dataset | Severe psoriasis (% of psoriasis) | Total psoriasis (% of population) | Non-psoriasis | HQ variants | $\lambda_{GWAS}$ |
|---|---|---|---|---|---|
| Estonian Biobank | 1 639 (11.6) | 14 167 (6.9) | 191 093 | 8 613 342 | 1.025 |
| FinnGen | 5 365 (42.2) | 12 708 (2.7) | 460 973 | 9 295 030 | 1.031 |
| HUNT | 1 491 (17.3) | 8 603 (10.0) | 77 802 | 7 582 614 | 1.024 |
| UK Biobank | 1 243 (13.2) | 9 426 (2.1) | 449 868 | 9 613 043 | 1.000 |
| Meta-analysis | 9 738 (21.7) | 44 904 (3.7) | 1 179 736 | 6 544 261 | 1.026 |

**Supplementary Table 2**: Statistical comparison of $PRS_{full}$ distribution between UK Biobank populations and BSTOP. P(difference between UKB and BSTOP mean): P-value for comparison of UK Biobank and BSTOP distributions using a Welch 2-sample t-test.

| **PRS** | **UK Biobank (sub)cohort** | **Proportion of BSTOP above 95th PRS percentile** | **Proportion of UKB-severe above 95th PRS percentile** | **P(difference between UKB and BSTOP mean)** |
|---|---|---|---|---|
| $PRS_{full}$ | UKB full | 34.2% | 21.6% | $<1\times10^{-300}$ |
| $PRS_{full\text{-}noHLA}$ | UKB full | 26.9% | 17.4% | $<1\times10^{-300}$ |
| $PRS_{full}$ | UKB psoriasis | 15.5% | 9.4% | $2.97\times10^{-300}$ |
| $PRS_{full\text{-}noHLA}$ | UKB psoriasis | 16.3% | 10.3% | $1.97\times10^{-245}$ |
| $PRS_{full}$ | UKB severe psoriasis | 6.4% | NA | $1.67\times10^{-37}$ |
| $PRS_{full\text{-}noHLA}$ | UKB severe psoriasis | 7.4% | NA | $3.48\times10^{-25}$ |

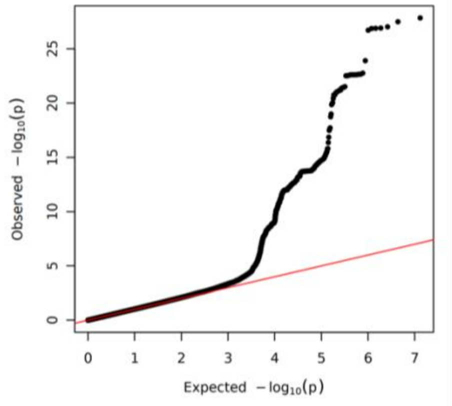


**Supplementary Figure 1**: Quantile-quantile plot for severe psoriasis GWAS meta-analysis of the four population-based cohorts.

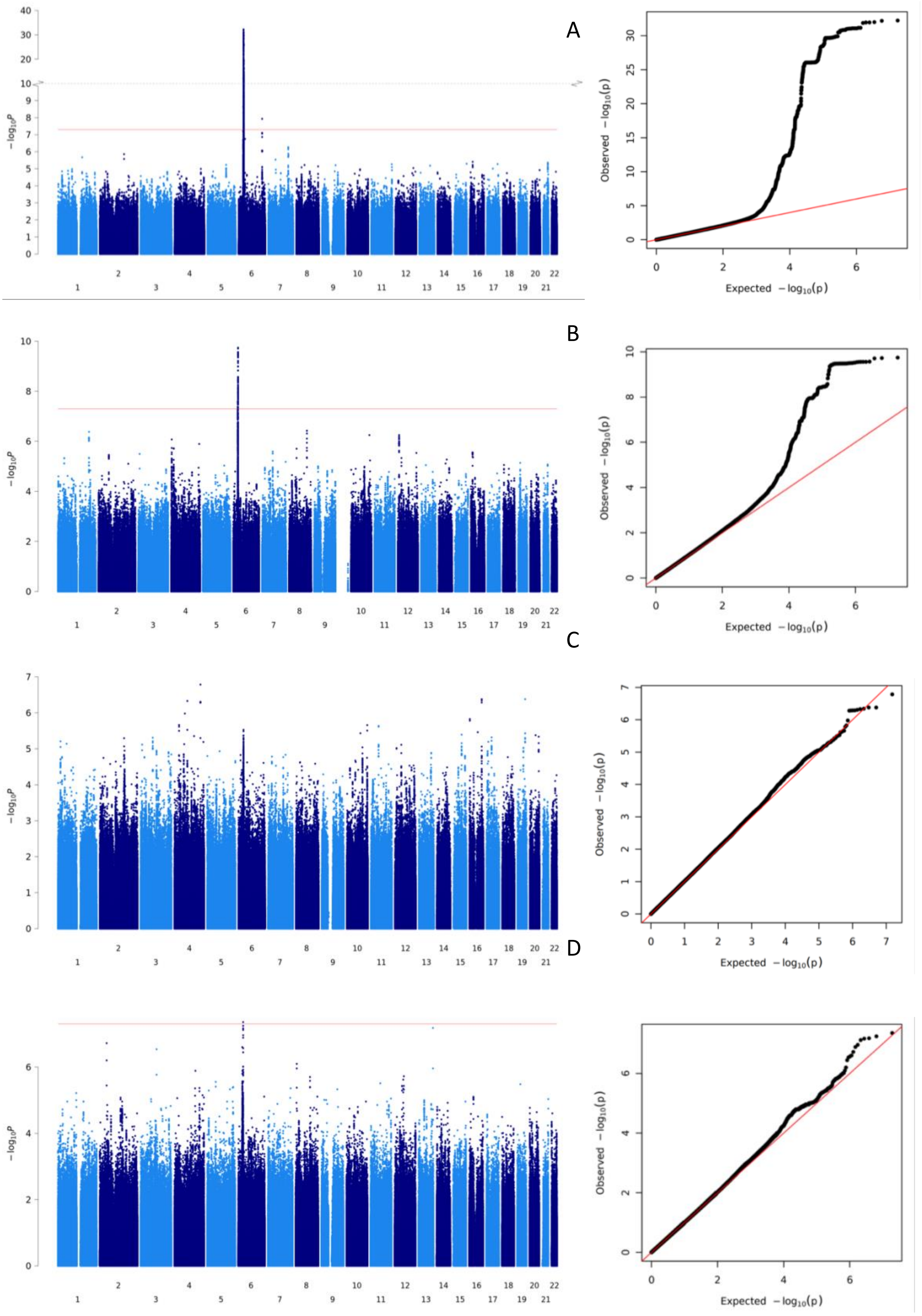
A
B
C
D
$-\log_{10}P$
Observed $-\log_{10}(p)$
Expected $-\log_{10}(p)$

**Supplementary Figure 2**: Manhattan and quantile-quantile plots for severe psoriasis GWAS of four population-based cohorts. Red line on Manhattan plots: $-\log(P) = 5 \times 10^{-8}$; Red line on QQ plots: x=y; A: Estonian Biobank; B: FinnGen; C: HUNT; D: UK Biobank.

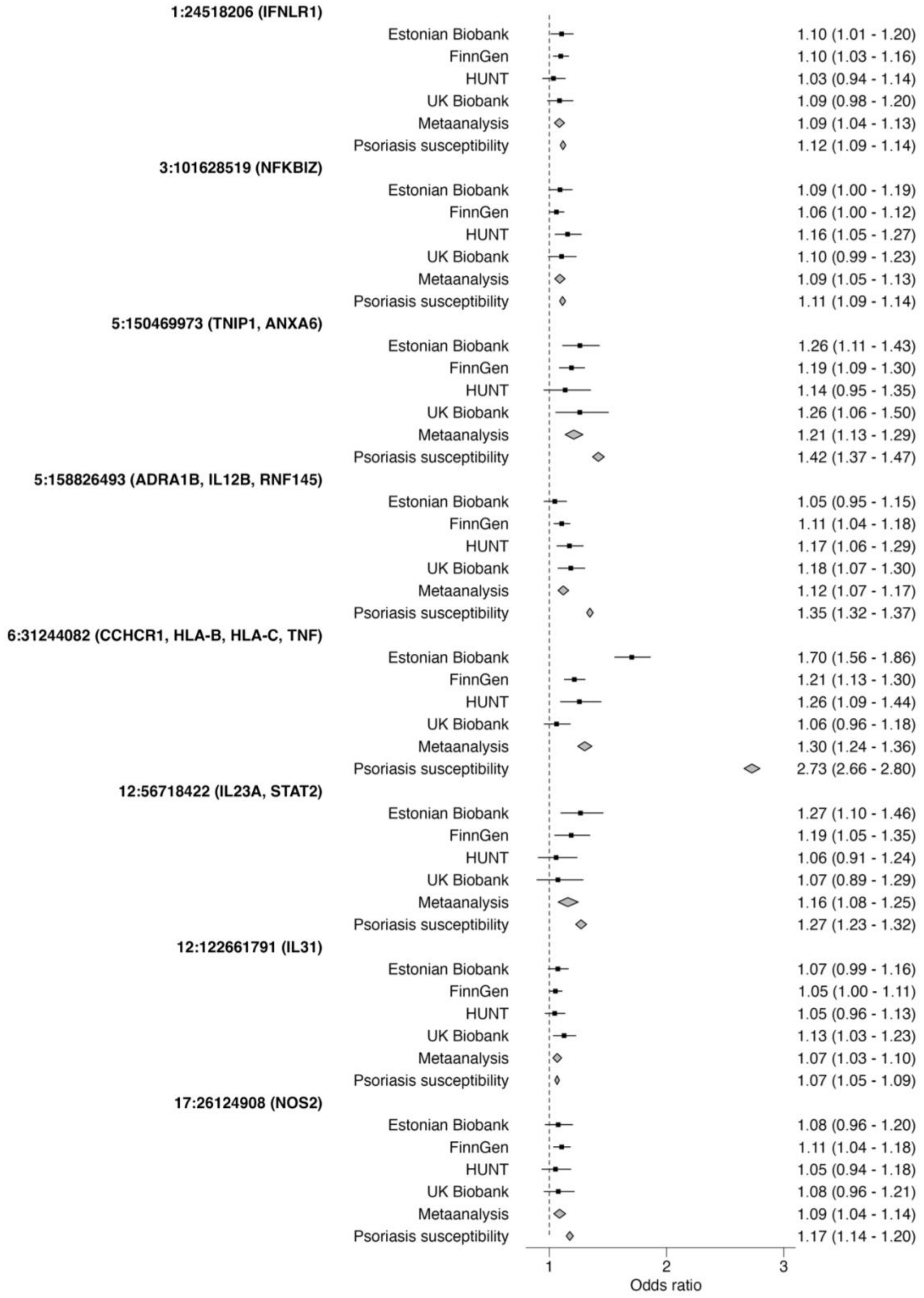


**Supplementary Figure 3**: Effect size estimates for eight psoriasis susceptibility SNPs significantly associated with severe disease after multiple testing (P<0.0005). Candidate genes are shown in brackets (Dand et al. 2023). Additive per-allele effect sizes presented numerically as: odds ratio (95% confidence interval).

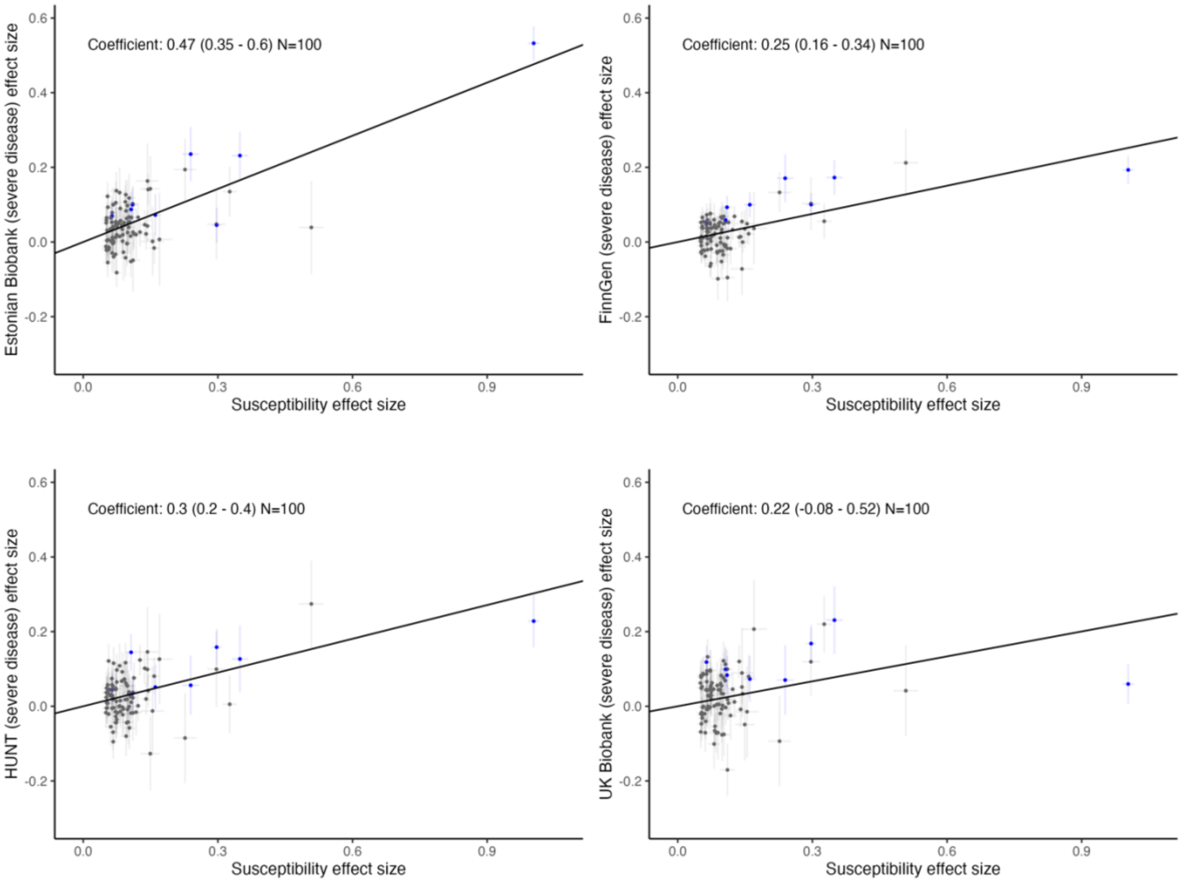


**Supplementary Figure 4**: Deming regression of severe psoriasis GWAS SNP effect sizes (betas) against psoriasis susceptibility effect sizes (betas), across four different population-based cohorts. SNPs with meta-analysis Bonferroni-corrected P-values < 0.05 are coloured blue.

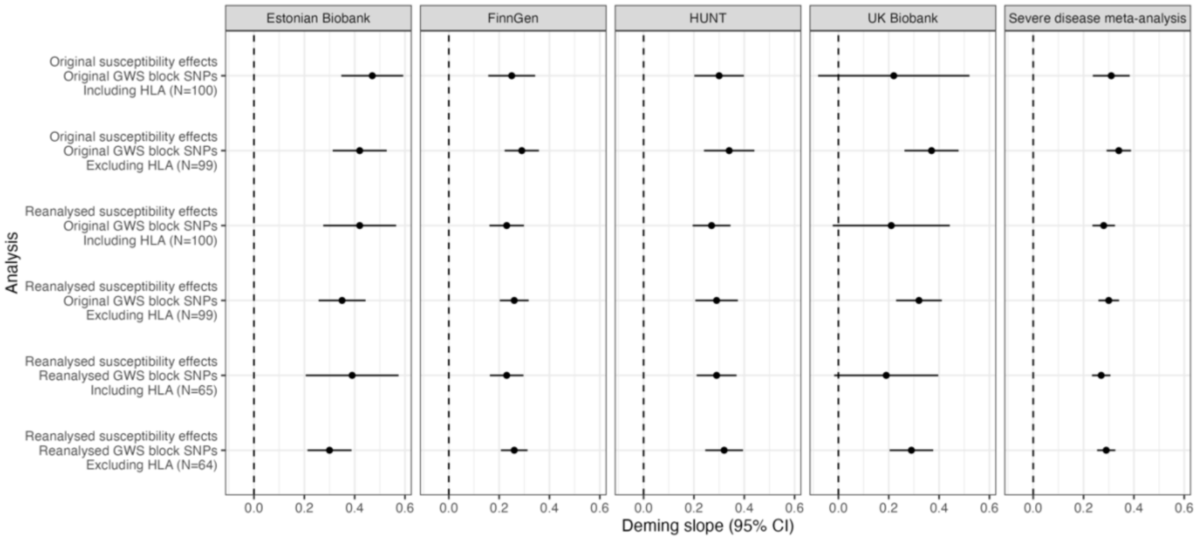


**Supplementary Figure 5**: Deming regression slopes comparing effect sizes of psoriasis susceptibility SNPs to effect sizes in severe disease for the meta-analysis and all four population-based cohorts individually. Sensitivity analyses where SNP selection and weighting have been adjusted to reflect complete independence from the population-based datasets are shown, as well as replication excluding the lead MHC SNP.

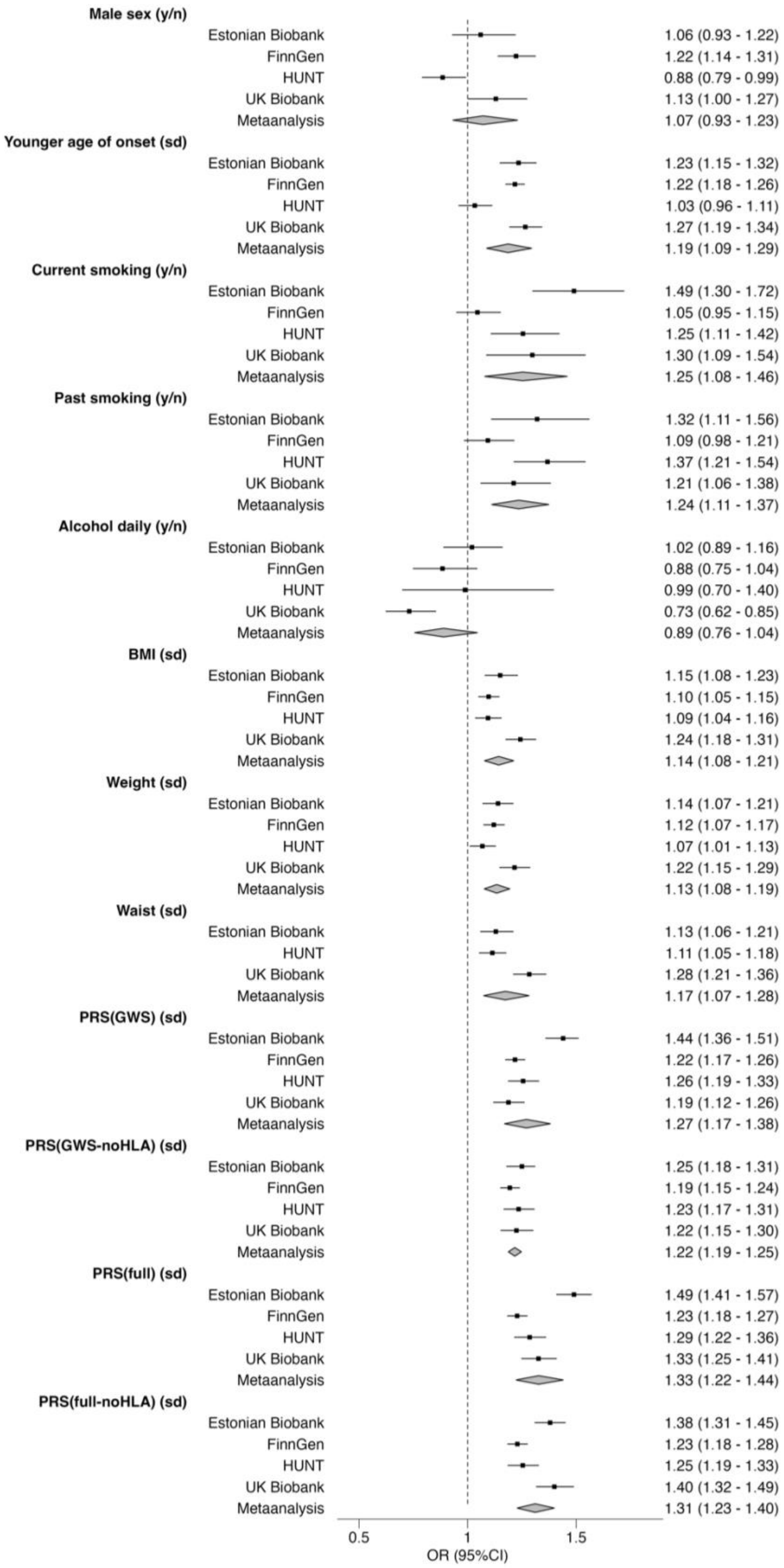


**Supplementary Figure 6:** Cohort-specific comparison of marginal effects of non-genetic factors and $PRS_{full}$ and $PRS_{GWS}$ with severe psoriasis. Effect estimates are derived from a meta-analysis of unadjusted logistic regression models comparing severe to non-severe psoriasis. y/n: effect size estimated for presence of exposure (“yes”) relative to absence (”no”); sd: effect size estimated per standard deviation change in continuous exposure within the psoriasis population; OR: Odds ratio. Effect sizes presented numerically as odds ratio (with 95% confidence intervals).

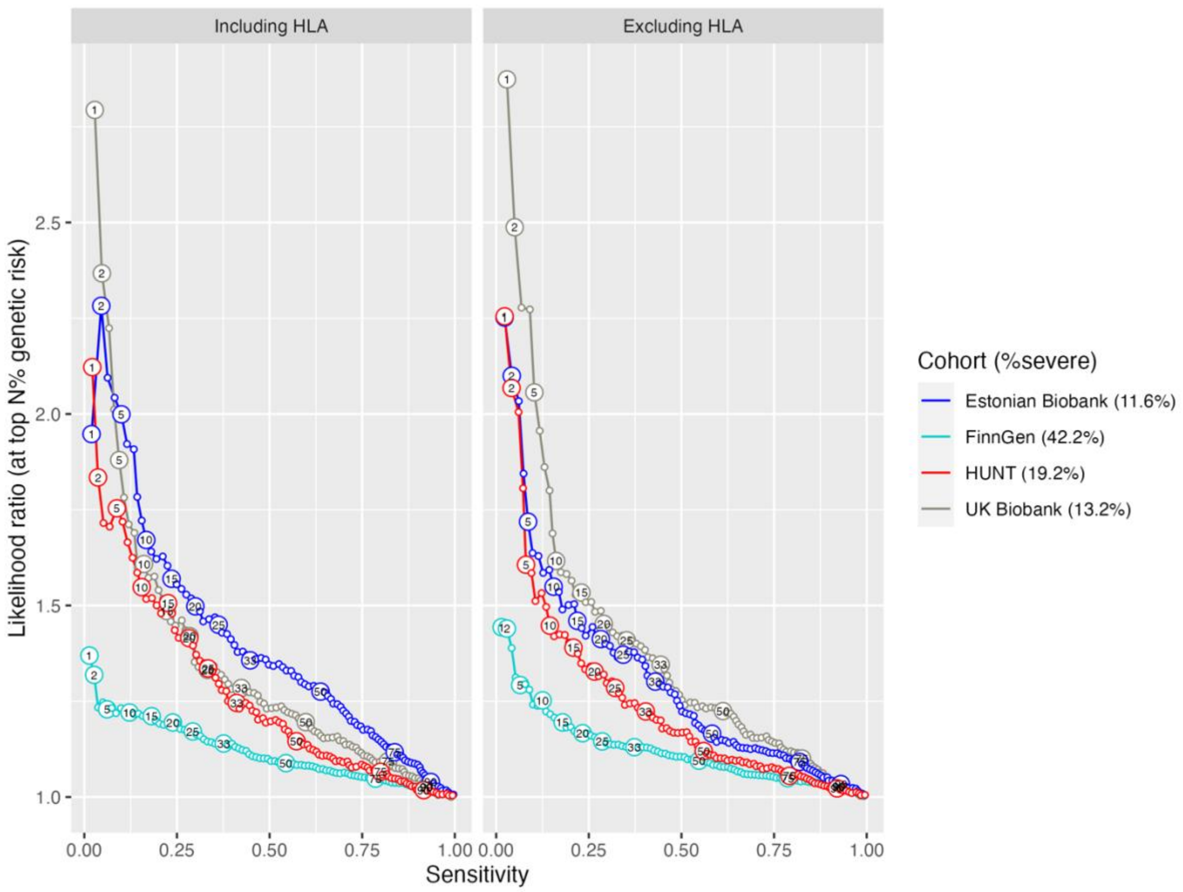


**Supplementary Figure 7**: Likelihood ratio plot against sensitivity, using top *N*% of psoriasis susceptibility $PRS_{full}$ as a “diagnostic test” in each of the European biobank cohorts. Top *N* percentage indicated as labels on selected points on the plot; *Sensitivity* = [number of severe individuals captured at PRS threshold] / [total severe individuals in cohort]; *Likelihood ratio* = [proportion of individuals captured by PRS who are severe] / [baseline rate of severe disease in cohort]

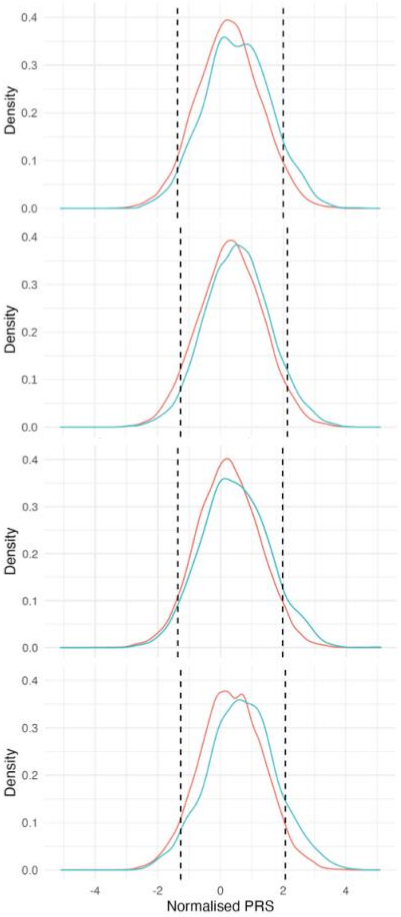


| Estonian Biobank | Full data | Bottom 5% | Middle 90% | Top 5% |
|---|---|---|---|---|
| Severe disease odds | 1:7.6 | 1:13.8 | 1:7.8 | 1:4 |
| Likelihood ratio | 1 | 0.59 | 0.98 | 1.72 |

| FinnGen | Full data | Bottom 5% | Middle 90% | Top 5% |
|---|---|---|---|---|
| Severe disease odds | 1:1.4 | 1:2.4 | 1:1.4 | 1:0.8 |
| Likelihood ratio | 1 | 0.70 | 1 | 1.29 |

| HUNT | Full data | Bottom 5% | Middle 90% | Top 5% |
|---|---|---|---|---|
| Severe disease odds | 1:4.8 | 1:6.2 | 1:4.9 | 1:2.6 |
| Likelihood ratio | 1 | 0.80 | 0.98 | 1.61 |

| UKB | Full data | Bottom 5% | Middle 90% | Top 5% |
|---|---|---|---|---|
| Severe disease odds | 1:6.6 | 1:11.4 | 1:6.9 | 1:2.7 |
| Likelihood ratio | 1 | 0.61 | 0.96 | 2.06 |

**Supplementary Figure 8**: Sensitivity analysis showing distributions and performance of susceptibility $PRS_{full\text{-}noHLA}$ in predicting severe psoriasis cases within psoriasis population in each biobank study. Cut-offs (dashed lines) displayed for individuals within the top 5%, middle 90% and bottom 5% of the (within-dataset) PRS distribution. Red line indicates PRS distribution of non-severe psoriasis population. Blue line indicates PRS distribution of severe psoriasis population. *Severe disease odds*: ratio of individuals with severe disease to individuals without severe disease. *Likelihood ratio*: ratio between the severe disease odds in each PRS group and the severe disease odds for all psoriasis cases.

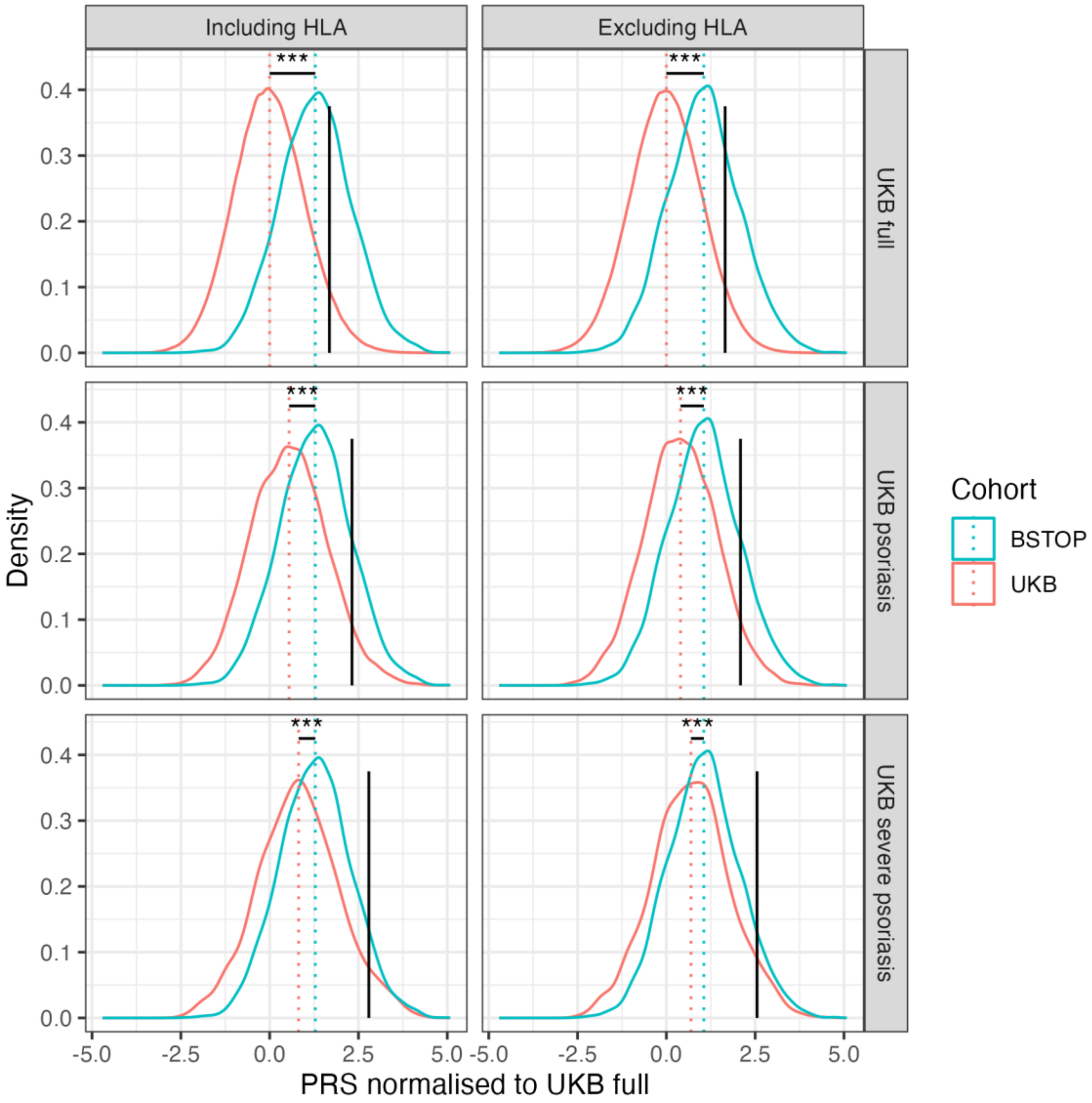


**Supplementary Figure 9**: Comparison of $PRS_{full}$ distributions between UK Biobank populations and BSTOP. Top row: all UK Biobank European ancestry participants (n = 459,294); middle row: all psoriasis cases in UK Biobank (n = 9,426); bottom row: severe psoriasis cases in UK Biobank (n = 1,243). Black vertical line indicates $95^{th}$ percentile of the PRS distribution in the UK Biobank participants in each row. ***: P-value<0.001 for difference between UKB and BSTOP PRS distribution means.

Supplementary Methods

#### Genotyping and GWAS

*Estonian Biobank*

The EstBB samples were genotyped at the Core Genotyping Lab of the Institute of Genomics, University of Tartu, using Illumina global screening arrays v1.0, v2.0 and v2.0_EST. Individuals with <95% call rate or sex mismatch between genetic sex and sex recorded in phenotype data were excluded from the analysis. Before imputation, variants were filtered by call rate < 95%, Hardy–Weinberg equilibrium P < 1 × 10-4 (autosomal variants only), and minor allele frequency < 1%. Prephasing was performed using Eagle v2.4.1 software, and imputation was performed using Beagle v.5.4 with the use of an Estonian population–specific imputation reference panel built from 2,297 whole-genome sequencing samples. Association testing was performed with a mixed-effects logistic regression model (REGENIE v3.0.3), including age, sex and ten principal components as covariates.

*FinnGen*

FinnGen (www.finngen.fi) is public-private research project that combines genomic data with Finnish health registry data. The project aims to genotype 500,000 Finnish individuals. FinnGen is a partnership of Finnish biobanks, their background organizations, pharmaceutical industry partners, and Finnish biobank cooperative (FINBB) (https://www.finngen.fi/en/partners). Data Freeze release 10, used in this analysis, contains 412,181 individuals with genotype and phenotype information.

Genotyping and quality control in FinnGen is described at [https://finngen.gitbook.io/documentation/methods/genotype-imputation/genotype-data](https://finngen.gitbook.io/documentation/methods/genotype-imputation/genotype-data). Briefly, samples were genotyping using Illumina (Illumina Inc., San Diego, CA, USA) and Affymetrix arrays (Thermo Fisher Scientific, Santa Clara, CA, USA). Samples were excluded based on the following criteria: no sex information or ambiguous sex, duplicates, >2% variant missingness, excess heterozygosity in common variants (MAF > 5%; +-3 standard deviation per batch). Samples with excess relatedness (($\hat{\pi}$>0.1) were also excluded in two rounds, first removing samples with > 500 related samples, then removing those remaining with > 50 counts. Variants with >2% missingness or Hardy-Weinberg equilibrium (HWE) P-value< $1x10^{-6}$ within a batch were removed. Variants were removed from all batches based on the following criteria: HWE P-value< $1x10^{-10}$ across all batches, missingness > 3% in any batch, or more than 15% of batches with missingness > 4%. Genotypes were pre-phased using Eagle 2.3.5 with the number of conditioning haplotypes set to 20,000. Genotype were subsequently imputed using the SISu v.4.0 reference panel using Beagle 4.1 (version 08Jun17.d8b). SISu 4.0 is a Finnish reference panel consisting of 8,554 whole genome sequences.

GWAS were performed using logistic mixed model implemented in REGENIE (v. 2.2.4) in the FinnGen sandbox environment. Both additive and dominant models were run, adjusting for age, sex, top 10 principal components, and genotyping batches. Specific genotyping batches were included as covariate if they were using in at least 10 cases or 10 controls. Variants with a minor allele count >= 5 were included in the analysis.

*HUNT*

The Trøndelag Health Study (HUNT) is a population-based cohort study carried out at four time points over approximately 40 years (HUNT1 [1984-1986], HUNT2 [1995-1997], HUNT3 [2006-2008] and HUNT4 [2017-2019]) (1,2). All inhabitants aged 20 years and over residing in Trøndelag County in Norway were invited to participate.

Participants from HUNT2-4 were genotyped using one of four different Illumina HumanCoreExome arrays (HumanCoreExome12 v1.0, HumanCoreExome12 v1.1, UM HUNT Biobank v1.0 and UM HUNT Biobank v2.0) (3). Genotype calling was performed with GenTrain v.2.0 in GenomeStudio v.2011.1 (Illumina). Samples with <99% genotype calls, with large chromosomal copy number variants, contamination >2.5% as estimated with BAF Regress (4), with genotypic and phenotypic sex discordance, and not of European ancestry were excluded, leaving 87,028 genotyped subjects. Genetic variants out of Hardy-Weinberg equilibrium (p-value <0.0001) were excluded. Samples were phased with Eagle2 v2.0.5 (https://alkesgroup.broadinstitute.org/Eagle/) and imputed with the positional Burrows-Wheeler transform (PBWT) v3.1 (https://github.com/richarddurbin/pbwt) (5).

GWAS was run in SAIGE v1.0.3 (6), using sex, birth year, genotyping batch and 10 ancestry principal components as covariates. Variants with MAF >3.2e-04 were included in the analyses, and dosages were used for imputed variants.

*UK Biobank*

The UK Biobank central team performed genotype calling and imputation. Genotyping was performed using the Affymetrix UK BiLEVE Axiom array (n ~50,000) and the Affymetrix UK Biobank Axiom array (n ~450,000) (7). Based on quality control metrics provided by UK

Biobank, we removed samples that exhibited sex mismatch, high relatedness (>3rd degree) to a large number (>200) of individuals, heterozygosity or missingness outliers, non-European individuals (according to kmeans cluster analysis) and withdrawals, leaving 462,817 individuals. Genome-wide imputation was performed by the UK Biobank central team using IMPUTE2 software and a reference panel derived from UK10K and 1,000 Genomes phase 3 haplotypes (8,9).

BOLT-LMM (10) was used to conduct genome-wide association testing, controlling for genotyping array type, sex, year of birth and 10 genotyping principal components. Effect sizes were approximated to the logistic scale using the formula: $\log(OR) = \beta / (\mu * (1 - \mu))$, where $\mu$ = case fraction. Standard errors were also converted by dividing by $(\mu * (1 - \mu))$.

*BSTOP*

BSTOP is an ongoing prospective observational study of patients with moderate–severe plaque psoriasis across >70 UK dermatology centres, which includes biological sample collection. Genome-wide genotyping array data were generated for BSTOP participants at the Institute of Psychiatry, Psychology and Neuroscience Genomics and Biomarker Facility at King’s College London (United Kingdom) using Illumina HumanOmniExpressExome-8, version 1.2, 1.3 and 1.6, and BeadChips (Illumina, San Diego, CA). Basic quality control and batch merging were performed using the process described in detail elsewhere (11,12).
